# The effects of Natalizumab Treatment on Astrocyte Metabolism in Multiple Sclerosis: A Longitudinal ^11^C-acetate PET study

**DOI:** 10.64898/2026.05.22.26353552

**Authors:** Hiroki Kato, Toru Koda, Hiroto Takahashi, Kenta Kurimoto, Makoto Kinoshita, Mikito Shimizu, Ryohei Yamamura, Naoshi Koizumi, Itsuki Sano, Yu Suzuki, Akihiro Tanaka, Kayako Isohashi, Noriyuki Tomiyama, Tatsusada Okuno

## Abstract

**Objective**

Astrocyte activation is increasingly recognized as an important component of multiple sclerosis (MS) pathology. Natalizumab (NTZ), a highly effective therapy for relapsing-remitting MS (RRMS), primarily blocks leukocyte trafficking into the central nervous system. However, its effects on astrocytic metabolism remain unclear. We investigated astrocyte-associated metabolic changes after NTZ treatment using quantitative 1-^11^C-acetate positron emission tomography (PET).

**Methods:** Seven patients with RRMS underwent quantitative 1-^11^C-acetate PET before and after NTZ treatment. PET-derived k2, an index of oxidative acetate metabolism, was analyzed voxel-wise and within GM and white-matter volumes of interest. Clinical status and brain magnetic resonance imaging (MRI) findings were assessed, and cognitive performance was evaluated using Rao’s Brief Repeatable Battery of Neuropsychological Tests.

**Results:** After NTZ treatment, k2 decreased in all patients compared with pretreatment levels. Both gray and white matter showed significant reductions, and voxel-based analysis demonstrated widespread decreases across cortical and subcortical regions of the cerebrum and cerebellum, with no regions showing significant posttreatment increases. MRI showed no worsening; Expanded Disability Status Scale scores were stable or improved, and cognitive performance was generally stable, with improvements in selected subtests.

**Interpretation:** Quantitative 1-^11^C-acetate PET demonstrated a whole-brain reduction in astrocyte-associated metabolism after NTZ treatment in RRMS, most prominently in gray matter. NTZ may modulate astrocyte activity, in addition to its established effects on peripheral immune cell trafficking.

## Introduction

Astrocyte activation is a key feature of inflammation in multiple sclerosis (MS) ^2–4^. During active MS, astrocytes are activated and become reactive astrocytes in response to stimulation by infiltrating immune cells and the release of pro-inflammatory cytokines^5^. Natalizumab (NTZ), a humanized monoclonal antibody against the integrin α4 subunit, is a pioneering, high-efficacy therapy for MS^6, 7^. Its primary mode of action is the specific blockade of α4β1 (VLA-4) and α4β7 integrins expressed on the surface of leukocytes ^7^. NTZ inhibits VLA-4 binding to its endothelial ligand, VCAM-1, thereby preventing leukocytes from firmly adhering to the vascular wall. This blockade almost eliminates the entry of new inflammatory cells into the central nervous system (CNS). This therapeutic effect is thought to result from immunosuppression caused by inhibition of T-cell trafficking across the endothelium^8–10^. In the cerebrospinal fluid, the number of lymphocytes, especially CD4^+^ T cells, is dramatically reduced after NTZ treatment^11^. Natalizumab also reduces microglial activity in MS^11^, its effects on astrocytes remain unclear. Acetate has been reported to be selectively taken up and metabolized by astrocytes in the central nervous system^12,13^. We have previously demonstrated, using qualitative and quantitative PET imaging, that 1-^11^C acetate uptake is significantly increased in patients with MS^14,15^. Thus, in this study, we aimed to investigate the effects of NTZ treatment on astrocyte metabolism in relapsing-remitting MS using PET imaging.

## Materials and methods

### Participants

Seven patients with RRMS (5 men and 2 women; mean age, 37.8 ± 7.6 years) who fulfilled the 2010 McDonald criteria^16^ and had experienced a clinical and/or radiological relapse within the previous year were prospectively enrolled (Table 1). The disease duration at enrollment was 80 ± 74 months, and the interval between the two PET scans was 30 ± 30 months. In Cases 1–5, only NTZ was administered during the two PET scans. In Case 6, glatiramer acetate was initiated 8 months before the initial PET scan and subsequently switched to NTZ because of worsening EDSS. In Case 7, dimethyl fumarate was initiated 29 months before the initial PET scan and was switched to NTZ owing to a new relapse. In Cases 6 and 7, NTZ was administered continuously for more than 2 years prior to the second PET scan. Cases 6 and 7 were also enrolled in our previously reported study^14^ without the results of the second PET scan images, which were captured in the current investigation. Participants provided written informed consent in accordance with the Declaration of Helsinki. This study was approved by the Ethics Committee of The University of Osaka Hospital.

**Table 1.** Clinical characteristics of the study participants.

| Variable | Pt 1 | Pt 2 | Pt 3 | Pt 4 | Pt 5 | Pt 6 | Pt 7 |
| --- | --- | --- | --- | --- | --- | --- | --- |
| Sex | M | F | F | M | M | M | F |
| Age group at PET1 | 50's | 40's | 30's | 40's | 20's | 30's | 40's |
| Disease duration, mo | 5 | 148 | 45 | 213 | 54 | 44 | 46 |
| Radiological relapse ≤1 yr before PET1 | Yes | Yes | Yes | Yes | Yes | Yes | Yes |
| Clinical relapse ≤1 yr before PET1 | Yes | Yes | Yes | Yes | No | Yes | Yes |
| Treatment at PET1 | None | GA | DMF | IFN-β | None | GA | DMF |
| Time from last relapse to PET1, mo | 5 | 1 | 1 | 6 | 53 | 13 | 1 |
| Treatment between PET scans | NTZ | NTZ | NTZ | NTZ | NTZ | GA 3 yr →<br>NTZ 2 yr | DMF 3 yr →<br>NTZ 2 yr |
| PET interval, mo | 9 | 8 | 4 | 10 | 11 | 62 | 63 |
| EDSS at PET1 | 4.5 | 1 | 2 | 2 | 1 | 2.5 | 5 |
| EDSS at PET2 | 4.5 | 1 | 2 | 2 | 0 | 2.5 | 5 |
| Clinical relapse after NTZ | No | No | No | No | No | No | No |
| MRI worsening after NTZ | No | No | No | No | No | No | No |

Abbreviations: Pt = patient; M = male; F = female; yr = years; mo = months; PET = positron emission tomography; EDSS = Expanded Disability Status Scale; MRI = magnetic resonance imaging; NTZ = natalizumab; GA = glatiramer acetate; IFN = interferon; DMF = dimethyl fumarate.

### MRI

Imaging was performed using a Philips Achieva dStream (Philips Healthcare, Netherlands) in two cases, a SIGNA Architect (GE Healthcare Japan, Tokyo) in one case, and a Discovery MR750 3.0T (GE Healthcare Japan, Tokyo) in the remaining cases. For 3D T1-weighted images, either an SPGR (spoiled gradient recalled) sequence (sagittal plane; slice thickness, 1.0 mm; matrix size, 256 × 256; in-plane resolution, 0.94 × 0.94 mm; field of view, 240 mm; repetition time, 7.10–7.20 ms; echo time, 2.74–3.30 ms; flip angle, 8°–11°) or an MPRAGE (magnetization prepared rapid acquisition with gradient echo) sequence (sagittal plane; slice thickness, 1.0 mm; matrix size, 256 × 256; in-plane resolution, 0.94 × 0.94 mm; field of view, 240 mm; repetition time, 2297.6 ms; echo time, 3.10 ms; flip angle, 8°) was used. Moreover, a FLAIR (fluid attenuation inversion recovery) sequence (sagittal plane; slice thickness, 1.4 mm; matrix size, 512 × 512; in-plane resolution, 0.47 × 0.47 mm; repetition time, 6000 ms; echo time, 91.1–372 ms; flip angle, 90°) was used. Diffusion tensor imaging (DTI) data were acquired using a single-shot echo-planar imaging sequence (sagittal plane; slice thickness, 2.6 mm; matrix size, 256 × 256; in-plane resolution, 0.94 × 0.94 mm; repetition time, 8500–15000 ms; echo time, 57.3–81.9 ms; flip angle, 90°; motion-probing gradient, 15 axes; b value, 1000 s/mm^2^). As a post-processing step for DTI, fractional anisotropy (FA) was calculated for each pixel.

### Synthesis of 1-^11^C-acetate

1-^11^C-acetate was synthesized by carbonation of a Grignard reagent, followed by acid hydrolysis. Specifically, 1-^11^C-acetate was obtained by reacting ^11^C carbon dioxide with methylmagnesium bromide, followed by hydrolysis with hydrochloric acid. The radiochemical purity of the tracer was > 98%.

### PET imaging

For all subjects, PET scans were performed in 3D acquisition mode using the Eminence SOPHIA SET-3000 BCT/X system (Shimadzu Corporation, Kyoto, Japan). Before the emission scan, transmission data were acquired with a rotating Cs-137 point source for attenuation correction. A radioactive tracer (400 ± 50 MBq) was injected into the precubital vein at a rate of 5.2 mL/min for 60 s, while a 60-min dynamic brain emission scan was initiated. Multi-frame tomographic images were reconstructed using the Dynamic Row-Action Maximum Likelihood Algorithm (DRAMA) with a 128×128 image matrix and a voxel size of 2.0 × 2.0 × 3.25 mm³. Each image dataset consisted of 35 frames (10 s × 9 frames, 15 s × 6 frames, 20 s × 3 frames, 30 s × 4 frames, 60 s × 4 frames, 180 s × 4 frames, 360 s × 3 frames, 600 s × 2 frames). The voxel values in the PET images were converted from cps/ml to Bq/ml based on the cross-calibration factor.

### Blood sampling

For input-function measurements, as in previous studies^15^, arterialized venous blood was collected instead of arterial blood to minimize invasiveness. A 22-gauge catheter was placed in the antecubital vein opposite the injection site for blood collection. Subsequently, the forearm was warmed with an electric blanket beginning 30 min before blood collection and continuing until the end of the scan. After warming, venous blood was collected to measure blood gas profiles.

Blood samples were collected at 20 s, 40 s, 60 s, 80 s, 100 s, 120 s, 150 s, 180 s, 210 s, 4 min, 5 min, 7 min, 10 min, 15 min, 20 min, 40 min, and 60 min after the start of the scan. The radioactivity and weight of whole blood or plasma obtained by centrifugation (3,000 rpm, 5 min) were measured using a well-scintillation counter (BSS, Shimadzu Corporation, Kyoto, Japan), and the radioactivity concentration (cps [counts per second]) of each blood sample was corrected for physical decay from the start of the scan. Blood data were corrected for metabolites using the following formula, which was derived and validated in previous studies:^17,18^

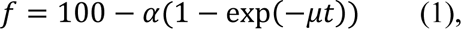

Here, f is the percentage of true 1-^11^C-acetate in plasma, with α = 91% and μ = 0.13/min^17^. Metabolite-corrected blood cell counts were converted from cps/g to Bq/ml using the cross-calibration coefficient.

### Quantitative analysis of PET

Similar to previous studies^15^, the metabolic rate of 1-^11^C-acetate in brain tissue was calculated using a one-tissue compartment model established in previous animal and human studies^18^. Briefly, the inflow of 1-^11^C-acetate (K1) and the clearance of total radioactive molecules produced by acetate metabolism in the brain (k2) were calculated. The clearance rate constant, k2, is thought to reflect the production of ^11^C CO_2_ and is therefore considered to be closely related to the oxidative metabolism of 1-^11^C -acetic acid^19^. K1, k2, and the volume of distribution (Vt) were calculated using the PXMOD module of PMOD 4.0 (PMOD Technologies LLC, Switzerland).

### Voxel-based statistical analysis

The summation-averaged images from all dynamic PET frames were coregistered to each subject’s T1-weighted 3D MR image using the PMOD 4.0 fusion tool. The k2 parametric images were also aligned to the magnetic resonance images using the same transformation matrix described above. T1-weighted 3D MRI images were anatomically segmented into gray matter, white matter, and cerebrospinal fluid probability maps using a segmentation tool based on an extension of the old unified segmentation algorithm ^20^ in Statistical Parametric Mapping 12 (SPM12) software (UCL, UK). Simultaneously, anatomical normalization was performed using the “Unmodulated Normalized” option to maintain image density. The coregistered PET images were normalized using the same transformation matrix derived from the segmentation described above. After anatomical normalization, the skull components were removed from the k2 images. For statistical image analysis, the PET images were smoothed using an isotropic Gaussian kernel with a FWHM of 12 mm.

After preprocessing, statistical comparisons were performed using a “paired t-test” with SPM12, without global scaling of voxel values or explicit masking to limit the comparison region. In this study, the statistical power was insufficient owing to the small sample size. Therefore, the statistical data were corrected for multiple comparisons using cluster-level inference based on random field theory rather than voxel-level inference. For the FA images, anatomical normalization, skull stripping, and smoothing were performed as described above. Statistical comparisons were then carried out using a “paired t-test” in SPM12, with proportional scaling and explicit masking, using FA maps produced from the mean DTI template (https://www.nitrc.org), with voxel values > 0.3.

### Analysis by volume of interest

Mean k2 values in the gray and white matter segments were evaluated using gray and white matter masks obtained from the tissue probability maps described in the previous section. Because of the limited PET resolution, the volume of interest (VOI) was defined according to the following conditions to reduce spill-in/spill-out effects between adjacent segments:

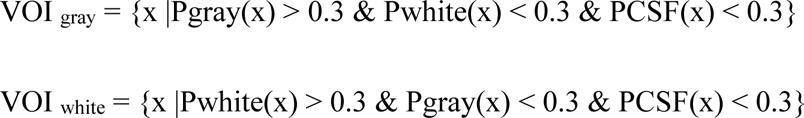

where Pgray(x), Pwhite(x), and PCSF(x) are the pixel values at voxel x in the image obtained after smoothing the segmentation masks of gray matter, white matter, and cerebrospinal fluid, respectively. Smoothing was performed using an isotropic Gaussian kernel with an FWHM of 8 mm, accounting for the PET spatial resolution.

### Cognitive function tests

Cognitive function was evaluated using Rao’s Brief Repeatable Battery of Neuropsychological Tests (BRBN-t)^21–23^. This battery consists of subtests assessing immediate and delayed verbal memory (Selective Reminding Test, SRT; SRT-Long-Term Storage, SRT-LTS; SRT–Consistent Long-Term Retrieval, SRT-CLTR; and SRT–Delayed Recall, SRT-D), immediate and delayed spatial memory (Spatial Recall Test, SPART; SPART–Delayed Recall, SPART-D), and sustained attention, concentration, and information processing speed (Paced Auditory Serial Addition Test at 3 s, PASAT3; 2 s, PASAT2; and Symbol Digit Modality Test, SDMT). Verbal fluency in response to semantic stimuli was assessed with Word List Generation (WLG). Cognitive function tests were administered immediately before each PET scan.

## Statistics

Cognitive function test scores and PET parameter values from the two PET imaging sessions were compared using the Wilcoxon signed-rank test. Statistical analysis was performed using SPSS 20 (IBM, USA).

## Results

### Disease activity before the first PET and outcomes after natalizumab treatment

All patients demonstrated evidence of disease activity during the year before the first PET scan; seven had radiologic relapses, and six also had clinical relapses (Table 1). At the time the first PET scans were performed, two patients were untreated, whereas the others were receiving either glatiramer acetate, dimethyl fumarate, or interferon-β (Table 1). After initiation of NTZ treatment, no patient exhibited evidence of disease activity until the second PET scan (NEDA-3; Table 1). The venous blood collected during PET imaging was sufficiently arterialized (Table 2).

**Table 2.** Arterialized venous blood gas values during PET acquisition.

| Variable | Mean (SD) |
| --- | --- |
| pH | 7.410 (0.019) |
| PaCO<sub>2</sub>, mmHg | 41.5 (3.6) |
| PaO<sub>2</sub>, mmHg | 106.1 (38.5) |
| HCO<sub>3</sub><sup>-</sup>, mmol/L | 26.3 (1.8) |
| Base excess, mmol/L | 1.6 (1.7) |
| O<sub>2</sub> saturation, % | 95.9 (5.3) |

Abbreviation: PET = positron emission tomography.

### Decreased whole-brain acetate metabolism after natalizumab treatment

Following NTZ treatment, k2 levels decreased in all cases compared with pretreatment levels (Figure 1). Pretreatment k2 levels were high in all cases relative to previously reported results for a healthy cohort, despite marked variation in the interval between the most recent relapse and acquisition of the first PET images (1–140 months; Table 1).

**Figure 1.**
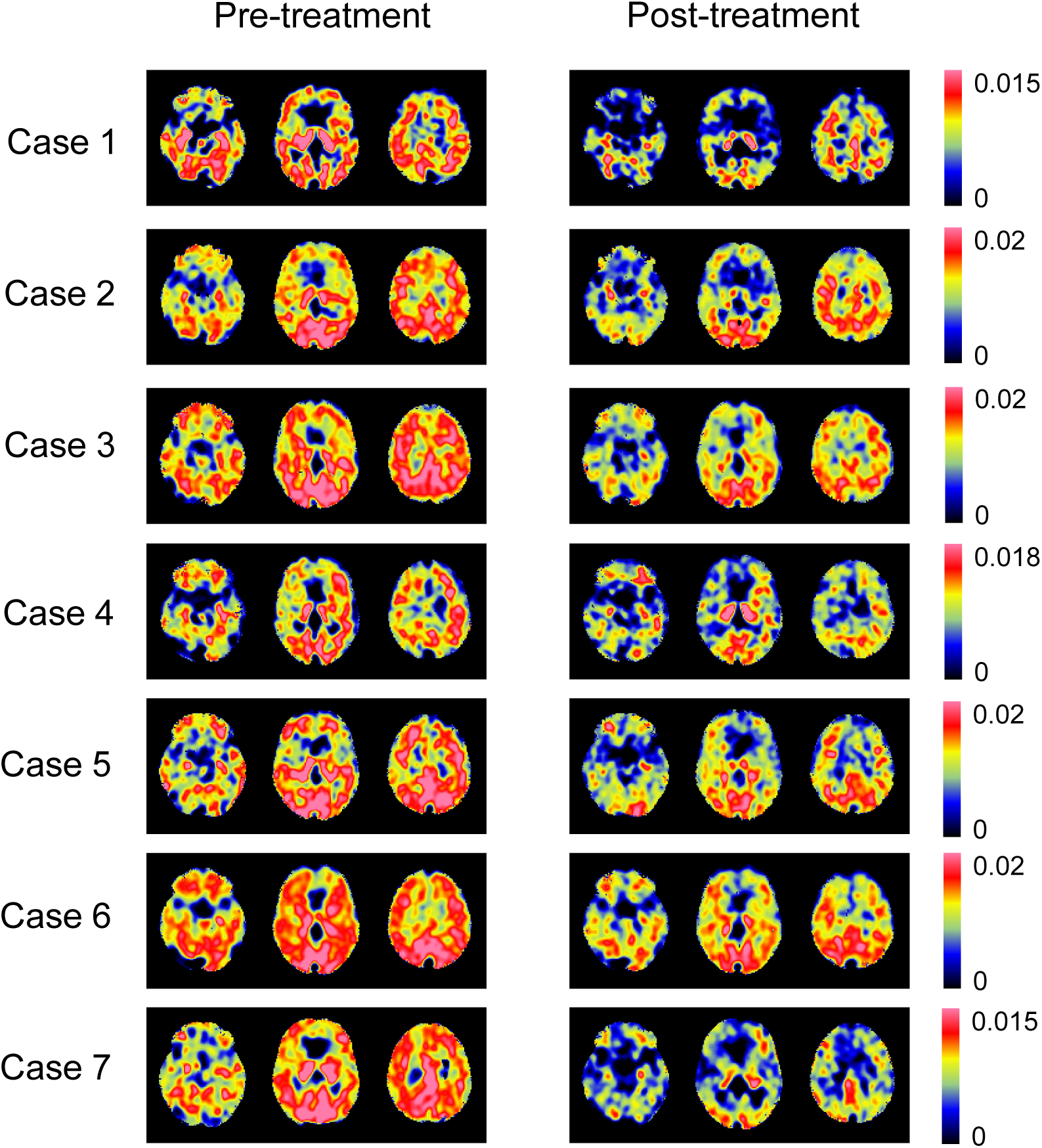
For cases 1–7, the k2 images were skull-stripped and anatomically normalized. The left column and right column present k2 images before and after NTZ treatment, respectively. The k2 values are represented by the color scale on the far-right side.

### Voxelwise changes in acetate metabolism and fractional anisotropy

Both gray- and white-matter segments showed a significant decrease in k2 levels (Figure 2). SPM analysis revealed a significant decrease in k2 levels after NTZ treatment in almost all cortical and subcortical regions of the cerebrum and cerebellum, including the thalamus. No brain region demonstrated significantly elevated k2 levels (Figure 3). Furthermore, although not statistically significant in the SPM analysis, the area with increased FA values after NTZ treatment was more apparent than the area with decreased FA values (Figure 4).

**Figure 2.**
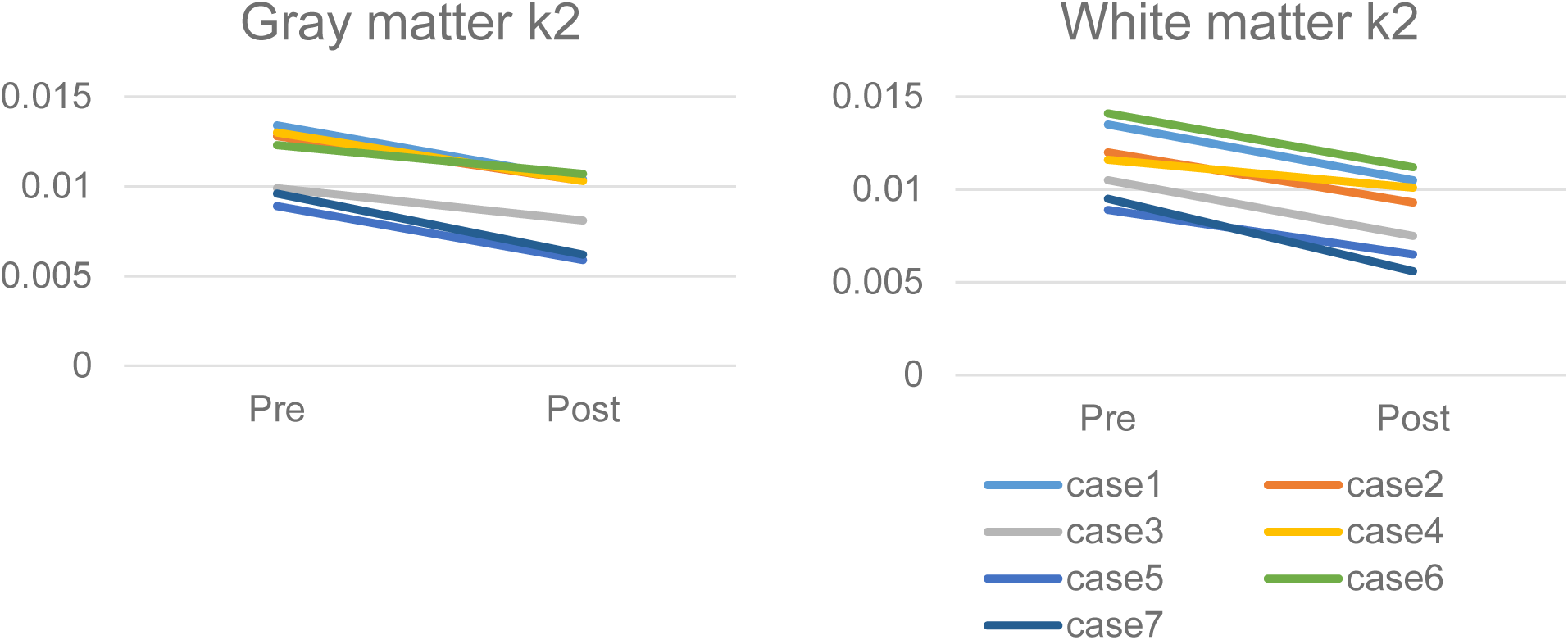
Using gray matter/white matter masks obtained from the segmentation of each patient’s MRI, the k2 values for gray matter (A) and white matter (B) were measured for the k2 images of each case.

**Figure 3.**
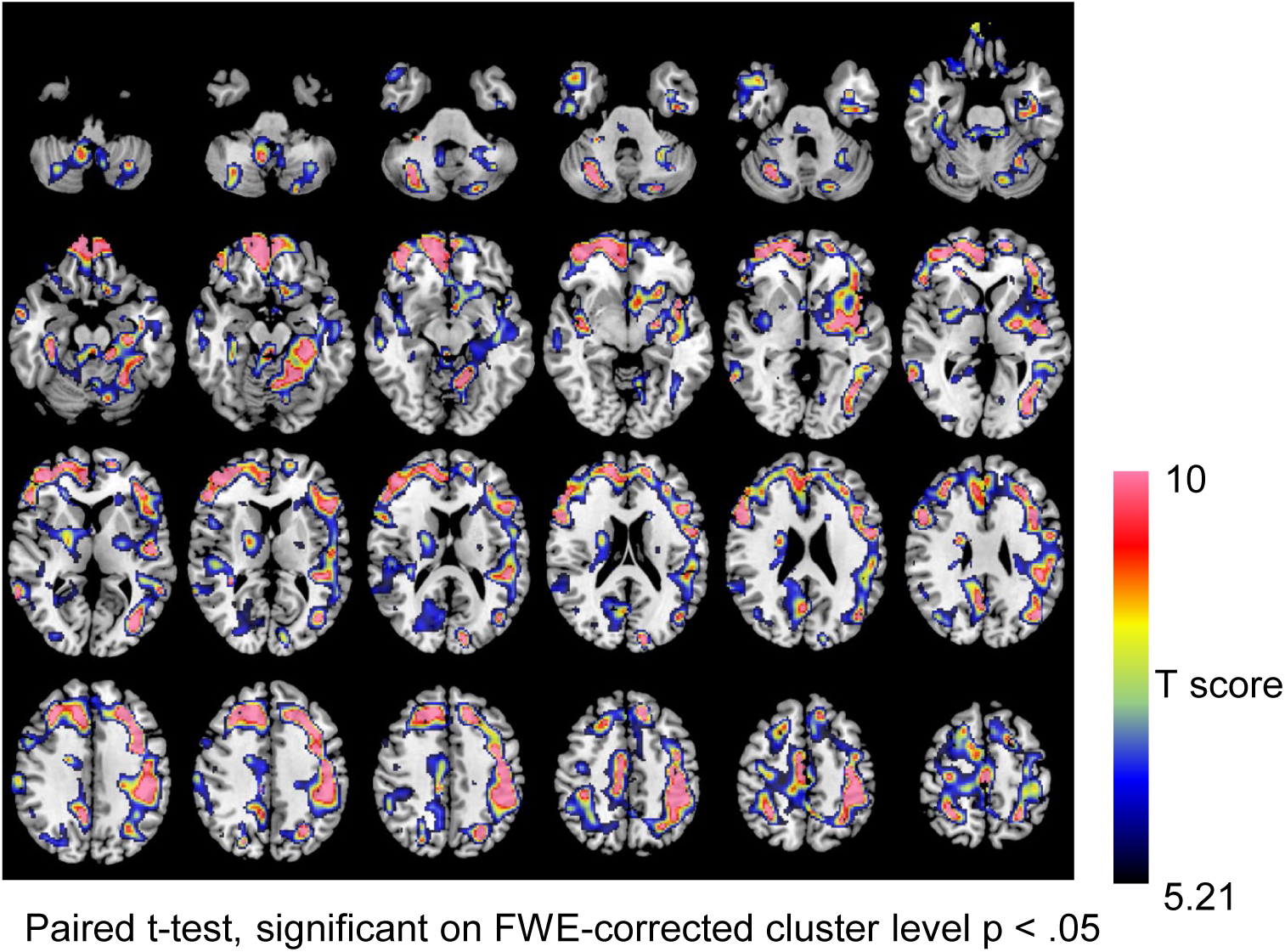
Statistical comparisons of k2 images before and after NTZ treatment in each case were performed using the “Paired t-test” in SPM12. The statistical data were corrected for multiple comparisons using cluster-level inference based on random field theory. The T-scores of voxels significant at the FWE-corrected cluster level are displayed on the MRI template using a color scale. k2 was significantly reduced after NTZ treatment in widespread cortical and subcortical regions of the cerebrum and cerebellum, including the thalamus.

**Figure 4.**
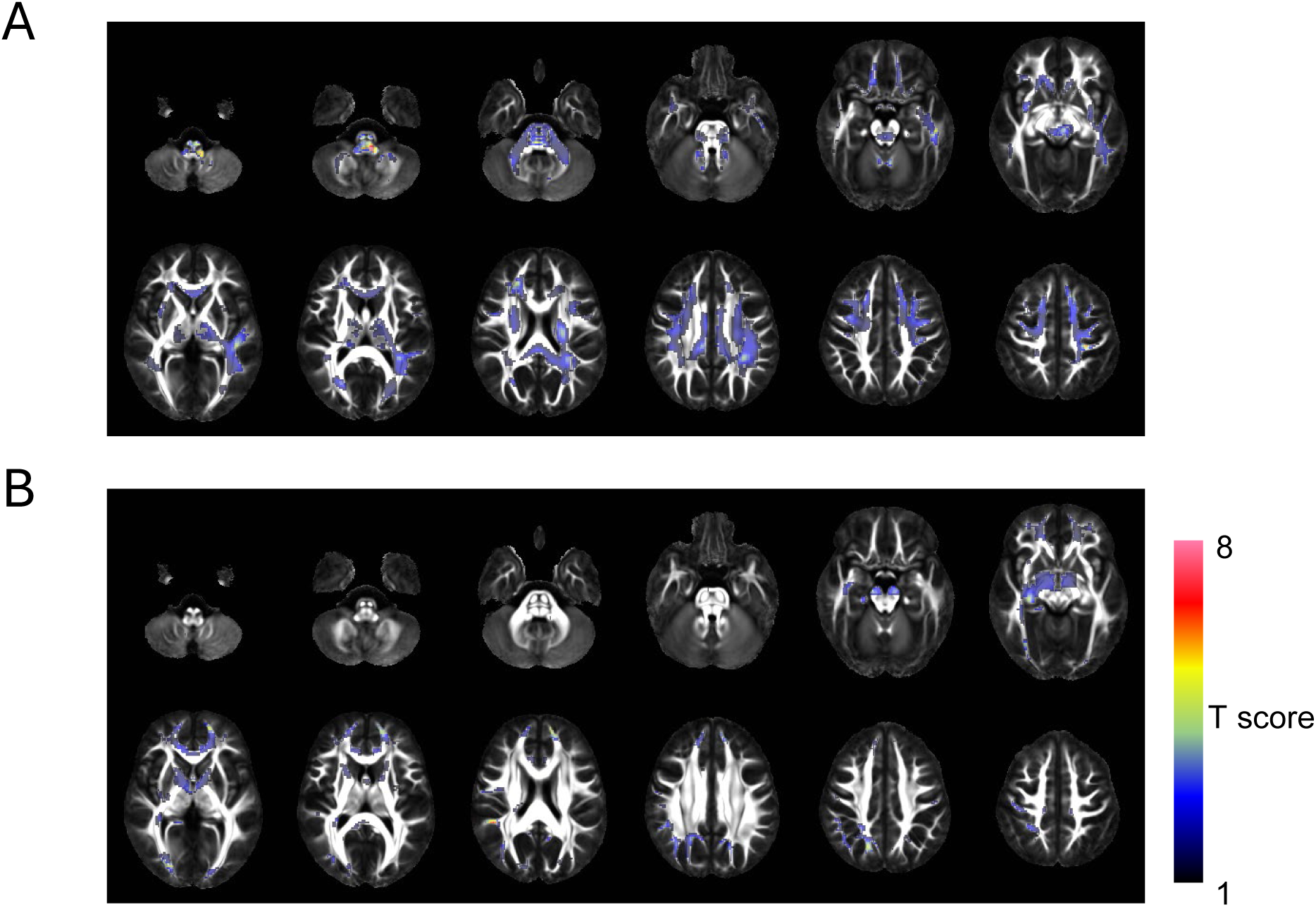
Statistical comparisons were performed using SPM12 and a “Paired t-test” on FA images from each case before and after NTZ treatment. For regions where FA values increased (A) and decreased (B) following NTZ treatment, the T-scores are displayed on a color scale overlaid on the FA template (IITmean_FA). The threshold for T-scores of voxels that were statistically significant (p < 0.001) without correction is 5.89.

### Disability and cognitive outcomes following natalizumab treatment

After NTZ treatment, EDSS scores remained unchanged in six patients and improved in 1 patient (case 5) (Table 1). On the BRBN-t, most subtests remained unchanged or showed a trend toward improvement. Significant improvements were observed in the SPART-D and WLG (Figure 5).

**Figure 5.**
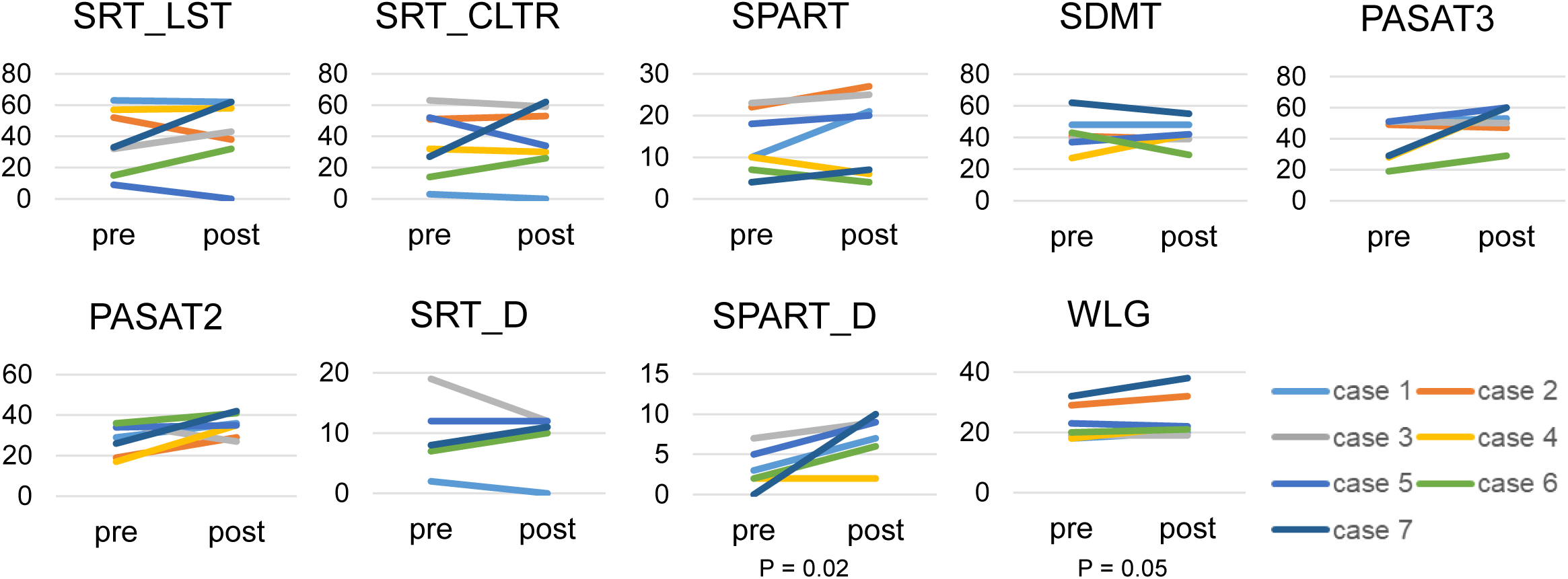
The results of cognitive function tests using Rao’s Brief Repeatable Battery of Neuropsychological Tests before and after NTZ treatment are displayed. Statistical comparisons were performed using the Wilcoxon signed-rank test. SRT: Selective Reminding Test, SRT_LTS: SRT–Long-Term Storage, SRT_CLTR: SRT–Consistent Long-Term Retrieval, SRT_D: SRT–Delayed Recall, SPART: Spatial Recall Test, SPART_D: SPART–Delayed Recall, PASAT3: Paced Auditory Serial Addition Test at 3 s, PASAT2: Paced Auditory Serial Addition Test at 2 s, SDMT: Symbol Digit Modality Test, WLG: Word List Generation

## Discussion

This study aimed to clarify the changes in astrocyte metabolism induced by NTZ treatment in RRMS using 1-^11^C-acetate PET, an imaging modality previously shown to primarily represent astrocyte metabolism. In all seven cases, k2 levels decreased throughout the brain after NTZ treatment. This change was particularly significant in cortical and subcortical regions. The k2 levels after NTZ treatment were considered close to the values previously reported in a healthy cohort^15^. None of the patients experienced clinical relapse, worsening MRI findings, or disability progression after NTZ initiation and before the second PET scan, and cognitive performance remained stable or improved in the selected domains^24^

MS is a chronic autoimmune demyelinating disease of the CNS, and its pathophysiology is understood as involving an attack by the peripheral immune system on the myelin sheath in the brain and spinal cord^23^. Nonetheless, recent advances in neuroimmunology have revealed that, beyond peripheral lymphocyte infiltration, glial cells, particularly astrocytes, are critical in disease progression and tissue repair ^24^.

Astrocytes are important in maintaining brain homeostasis, supporting neuronal metabolism, and regulating synaptic function. In MS, some of them are thought to transform into “reactive astrocytes,” which are believed to contribute to disease progression or repair ^24^. Reactivity of astrocytes can be assessed by measuring their metabolism ^17,18^. Using 1-¹¹C-acetate PET, we demonstrated both qualitatively^13^ and quantitatively^14^ that astrocyte metabolism is elevated in MS compared with healthy individuals. Astrocytes form the glia limitans with their foot processes, thereby maintaining the structural integrity of the blood-brain barrier (BBB)^25^. In MS, the BBB is functionally broken down, allowing peripheral immune cells to enter the CNS. Astrocytes are closely involved in neurovascular and inflammatory processes in MS and may contribute to interactions between the BBB and infiltrating immune cells.

NTZ is a humanized monoclonal antibody targeting the integrin α4 subunit and is a pioneer among highly effective therapies for multiple sclerosis. Its primary mechanism of action involves specific blockade of the α4β1 (VLA-4) and α4β7 integrins expressed on white blood cells ^7^. This blockade almost completely prevents the influx of new inflammatory cells into the CNS. Large-scale clinical trials, such as the AFFIRM trial, have demonstrated that gadolinium-enhancing lesions on MRI disappear early after treatment initiation and that the relapse rate is reduced by approximately 68% ^25^. Glial fibrillary acidic protein (GFAP) is used as a marker of astrocyte activation or damage. At baseline, serum GFAP (sGFAP) levels are significantly elevated in patients with MS compared with healthy controls, reflecting persistent astrocyte reactivity (astrogliosis) ^26^. Sainz-Amo et al. reported that, in patients who showed no symptom recurrence and whose disease progression had halted on MRI, sGFAP levels decreased significantly within 6 months of treatment initiation. This suggests that, by blocking leukocyte infiltration, astrocytes within the CNS are relieved of inflammatory stimuli and transition to a state closer to quiescence. Conversely, in patients whose disease activity persisted despite treatment, no decrease in sGFAP levels was observed ^26^. Here, we evaluated astrocytic activity from a metabolic perspective, and our results are consistent with those of Sainz-Amo et al.

Recently, the lymphocyte-microglia-astrocyte axis has been proposed as a conceptual framework to comprehensively understand how peripheral immune cells infiltrate the central nervous system (CNS) and subsequently activate resident glial cells in the pathophysiology of MS^26^. NTZ treatment is thought to affect the lymphocyte-microglia-astrocyte axis through the following mechanism: infiltrating T cells release effector cytokines, such as interferon-gamma and interleukin-17 ^27^. These cytokines bind to receptors on microglial surfaces, inducing them to enter an activated state^28^. Activated microglia then secrete additional inflammatory mediators, which activate astrocytes^28^. Natalizumab suppresses the activation of astrocytes and microglia, which are resident immune cells. *In vivo* studies using 18-kDa translocator protein (TSPO)-PET have confirmed that, 1 year after natalizumab treatment, microglial activation is significantly reduced in the normal-appearing white matter and at the rim of chronic lesions^1^.

Astrocytes and microglia engage in “cellular crosstalk,” by mutually activating each other through cytokines and chemokines; this may modulate astrocyte activity^5^. Results from Acetate PET and TSPO-PET studies support the notion that diffuse smoldering neuroinflammation associated with the progression of multiple sclerosis (MS)^29^ can be therapeutically targeted using NTZ^1^.

Our previous studies have demonstrated that, in patients with MS, metabolism in both gray matter and white matter is elevated compared with that in healthy controls and that pathological changes are significantly greater in the white matter ^14,15^. Furthermore, statistical image analysis indicates that the pathological elevation of acetate metabolism is particularly pronounced in the white matter, and a link to axonal damage has been suggested^15^. This study revealed that NTZ administration reduced acetate metabolism in both the gray and white matter. Statistical analyses revealed significant treatment-related changes in the cortex and the thalamus. This may be because, unlike pathological changes, the suppression of inflammation induced by drug therapy extends throughout the brain.

We demonstrated decreased FA levels in patients with MS^15^. Although the interpretation of changes in FA remains under debate, a decrease in FA has been reported to reflect demyelination and/or axonal damage^30,31^. The results of this study indicate that NTZ treatment maintained axonal function. CSF neurofilament light chain levels, a highly sensitive marker of axonal damage, decreased in correlation with the reduction in inflammation-related markers observed with natalizumab treatment^32^. These results support those findings.

An open-label, multicenter study (the Evaluation of Natalizumab for the Relief of MS-Associated Fatigue (ENER-G) study) demonstrated that cognitive performance improved or remained stable over 48 weeks after natalizumab initiation^33^. Furthermore, in a study using a specific neuropsychological battery, 52.5% of patients showed improvement in cognitive function within 1 year of starting treatment, 30.0% remained stable, and only 17.5% showed deterioration^34^. Improvements in PASAT scores in the Natalizumab Safety and Efficacy in Relapsing-Remitting Multiple Sclerosis (AFFIRM) trial suggested neuropsychological benefits compared with placebo^35^. In this study, none of the BRBN-t subscales showed deterioration following NTZ treatment; significant improvements were observed in the SPART–D, a test of visuospatial episodic memory, and the WLG, a verbal fluency task. A study that reanalyzed MRI data from the AFFIRM trial using the latest deep-learning techniques found that NTZ reduced the decline in gray matter volume by 64.3% over 2 years compared with placebo^36^.

Specifically, the effect of inhibiting thalamic atrophy, which is considered the “hub of cognitive function” in MS, was significant, with a 57.0% reduction in atrophy observed compared with placebo by year 2 of treatment^36^. The cognition-enhancing effects of NTZ treatment may be related to significant improvements in astrocyte metabolism in the cerebral cortex and thalamus.

Several limitations should be acknowledged. First, the sample size was small. Second, this was a longitudinal within-subject study without a contemporaneous control group. Third, two of the seven patients had received a different drug before NTZ. These treatments are believed to alter astrocytic metabolism. Both patients had been switched to NTZ because of insufficient disease control and had received NTZ continuously for more than 2 years before the second PET scan. Therefore, although the effects of prior therapies cannot be fully excluded, they are unlikely to account entirely for the consistent direction of change observed across all patients.

In conclusion, quantitative 1-^11^C-acetate PET revealed that NTZ treatment in RRMS resulted in a whole-brain decrease and normalization of astrocyte metabolism relative to pretreatment levels. Furthermore, ¹¹C-acetate PET may enable the detection of T-cell-mediated silent inflammation in the central nervous system by assessing astrocyte metabolic changes and may serve as a useful tool for monitoring treatment efficacy.

## Acknowledgements

This study was supported by JSPS KAKENHI (grant numbers 25K10770, 23K27517, and 25H01863), JST ERATO (grant number JPMJER2102), and by AMED (grant number 23gm1810003h0002). This research was conducted using the REDCap system operated by the Osaka University Graduate School of Medicine and the Osaka University Hospital. We acknowledge the assistance of Chika Fukutomi and the entire staff of the Department of Nuclear Medicine, Osaka University Graduate School of Medicine, with PET data acquisition and subject care.

## Author contributions

TO and HK conceived the project and designed the overall study. HK and TK prepared the manuscript and figures/tables. HK and HT analyzed all 1-¹¹C-acetate PET and brain MRI data. KK, RY, NK, IS, AT, KI, and contributed to the acquisition of 1-¹¹C-acetate PET data. TK and TO performed clinical assessments. All authors critically reviewed the manuscript for important intellectual content and approved the final version.

## Data Availability

The dataset for this study is available from the corresponding author on reasonable request.

